# Assessment of Added Activity of An Antitumor Agent

**DOI:** 10.1101/2022.10.14.22281099

**Authors:** Cong Chen, Linda Zhiping Sun, Yixin Ren, Eric H. Rubin, David M. Weinstock, Emmett V. Schmidt

## Abstract

An unprecedented number of novel oncology drugs are under preclinical and clinical development, and nearly all are developed in combinations. With an over-reliance on biological hypotheses, there is less effort to establish single agent activity before initiating late clinical development. This may be contributing to a decreased success rate going from phase 1 to approval in the immunotherapy era. Growing evidence in clinical trial data shows that the treatment benefit from most approved combination therapies can be explained by the independent drug action model. Using this working model, we develop a simple index to measure the added antitumor activity of a new drug based on mean response duration, an endpoint that naturally combines both response status and duration information for all patients, which is shown to be highly predictive of clinical benefit of FDA-approved anti-PD-(L)1 immunotherapies. This index sheds light on challenges and opportunities in contemporary oncology drug development and provides a practical tool to assist with decision-making.

## Introduction

Combination therapy is an important paradigm in oncology treatment that addresses the heterogeneous biology within tumors. It is of utmost interest to assess the added antitumor activity (AAA) of a new drug when it is combined with the standard-of-care (SOC) or any other therapy. Antitumor activity is normally assessed with objective response rate (ORR) and duration of response (DoR) in Phase 1/2 clinical trials, often with single-arm cohorts. Progression-free-survival (PFS) is limited to randomized Phase 2 trials because it is confounded with natural history of the disease. As immune checkpoint inhibitors, which result in prolonged responses among a subset of patients, became a backbone in cancer treatment over the last decade, there has been growing interest in durable response. However, when isolated from ORR, DoR can be misleading as it is entirely based on responders and ignores non-responders^1^. A natural remedy is to consider the mathematical product of the two (ORR x DoR). The product measures the mean response duration (MRD) or AUC of response in all patients, whereas DoR for non-responders is technically zero, and it can be formally estimated when patient-level data is available^2-4^. Restricted to trial follow-up time, the AUC was found to outperform ORR and PFS in predicting survival benefit in an extensive simulation study based on Phase 3 trial data of pembrolizumab in melanoma^5^. The superiority of MRD is confirmed in an analysis of published Phase 3 trial data of all anti-PD-(L)1 immune checkpoint inhibitors approved by the US FDA in first-line advanced/metastatic tumors (Figure 1). We focus on MRD for AAA assessment due to its superior performance as well as its broad applicability to both randomized and single-arm trials.

**Figure 1.**
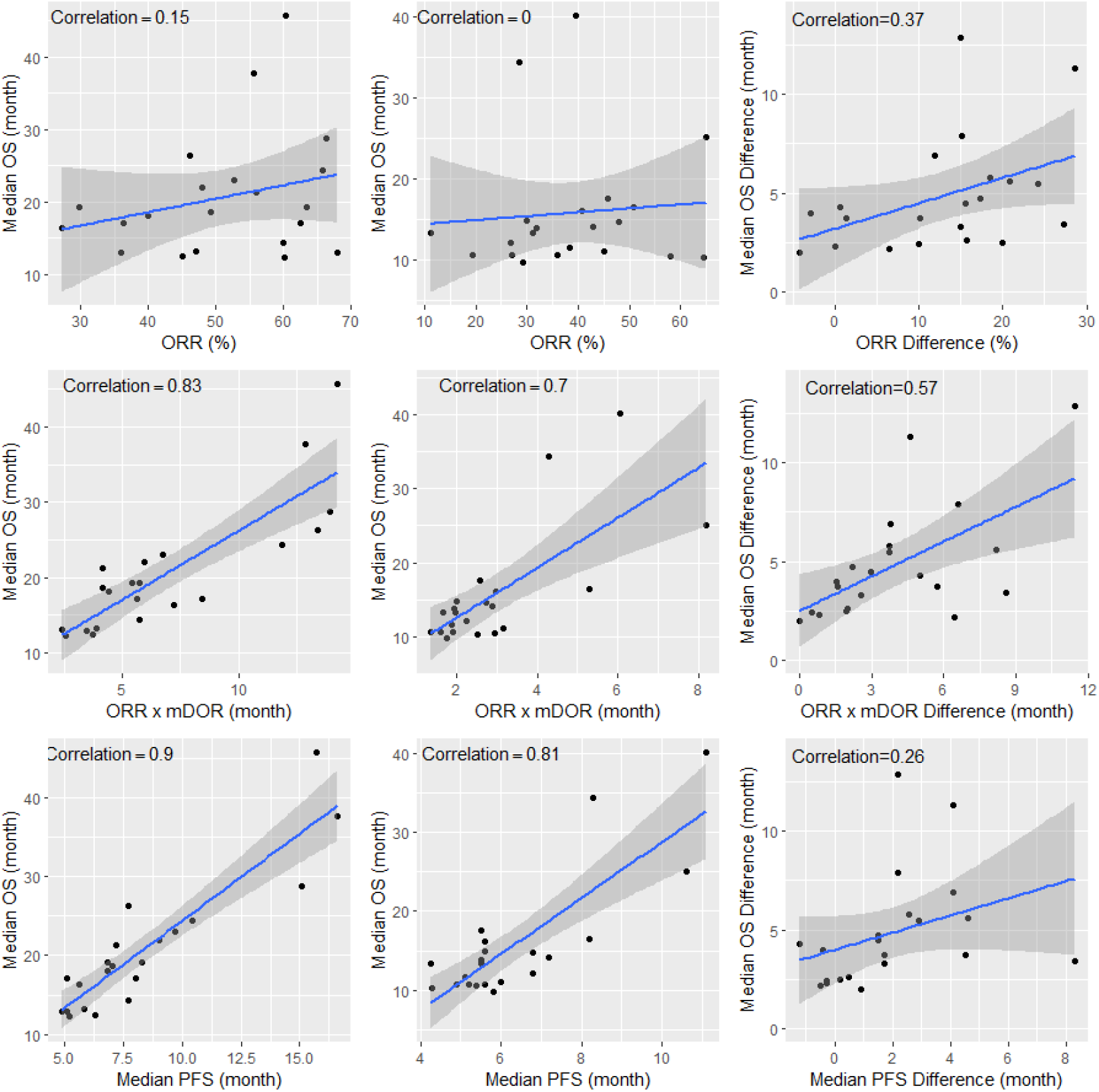
Comparison between ORR (top), MRD (middle) and PFS (bottom) in terms of correlations with median OS based on published Phase 3 trial data of all anti-PD-(L)1 immune checkpoint inhibitors approved by FDA in first-line advanced/metastatic tumors either as a monotherapy or a combination therapy (Left panel: experimental arm; Center panel: control arm; Right panel: difference between experimental arm and control arm). The blue line represents the regression slope, and the shaded area represents the 95% confidence interval (the narrower the area the higher the correlation). Only those trials referenced in the US labels as of July 1, 2022, with complete ORR, median DoR, median PFS and median OS data are included in the analysis (Supplementary Appendix). MRD difference has higher correlation with OS improvement than ORR or PFS difference. Besides, a positive OS difference may not be always associated with a positive ORR difference or a positive PFS difference, but often is with a positive MRD difference. Overall, MRD is a better early endpoint than ORR and PFS.

The independent drug action model will be used to assist with the assessment. Built upon patient-to-patient variability, the model assumes that the benefit a patient receives from a combination therapy is driven by the drug component the patient’s tumor is most sensitive to. In general, the putative benefit to the two components may be correlated. A positive correlation implies the possibility of cross-resistance, while a negative correlation, albeit less common, implies the possibility of a collateral sensitivity^6-9^. However, consistent with findings from preclinical models^10^, the benefits in early efficacy endpoints (ORR, PFS, DoR) for many combination therapies have been successfully predicted under the assumption of zero correlation or a small positive correlation^6-9,11-13^. We will provide a simple AAA index of MRD under the assumption of zero correlation for practical purpose. To pave the way, we will illustrate the application of independent drug action model to ORR and PFS first.

## Methods

### AAA of ORR

When Drug 1 and Drug 2 act independently, the ORR of the combination therapy is expected to be around *p*_1_ + *p*_2_ − *p*_1_*p*_2_, where *p*_1_ and *p*_2_ are the respective ORRs of the two drug components. This is widely known as the Bliss independence model^14^ (Figure 1). It was found that ORRs of anti-PD-(L)1 immune checkpoint inhibitor combination therapies reported in clinical trials largely fit the Bliss independence model prediction^11^. The Bliss independence model is mathematically equivalent to the assumption that a patient’s response to a combination therapy is the best response to a drug component (aka independent drug action model^6^). Taking out *p*_1_ from the model prediction, an AAA index of ORR from Drug 2 is clearly *p*_2_ − *p*_1_ *p*_2_ or *p*_2_(1 − *p*_1_). For example, when the ORRs for both drugs are as high as 50%, the ORR for the combination would be around 75% (i.e., 0.5+0.5-0.5*0.5) and the AAA from drug 2 would be only 25%. But when ORR for Drug 1 is reduced to 20%, the AAA from Drug 2 would be 40% despite lower ORR for the combination (60%). This analysis highlights the challenges in improving upon SOC with high efficacy, a commonly known phenomenon to drug developers.

### AAA of PFS

When the independent drug action model is applied to PFS, it is assumed that a patient’s PFS duration to a combination therapy is the best duration to a drug component^6-8^ (Figure 2). A mathematically equivalent form in terms of PFS rate at any time is derived from this assumption to facilitate the calculation of the Kaplan-Meier estimate of the predicted PFS as well as its confidence intervals^12^. PFS durations of the two drug components are assumed to follow an exponential distribution, a common working assumption in practice. In this case the PFS duration for the combination therapy is around 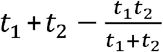, where *t*_1_ and *t*_2_ are the respective PFS durations for the two drug components (Supplementary Appendix). The AAA index of PFS from Drug 2 is around 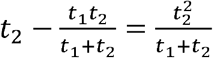. For example, when PFS duration is 3 months for Drug 1 and 6 months for Drug 2, PFS duration for the combination would be around 7 months and the AAA of PFS for Drug 2 would be around 4 months. But when PFS duration for Drug 1 is increased to 6 months, the AAA of PFS for Drug 2 would be reduced to around 3 months, again highlighting the challenges in improving upon SOC with high efficacy.

**Figure 2.**
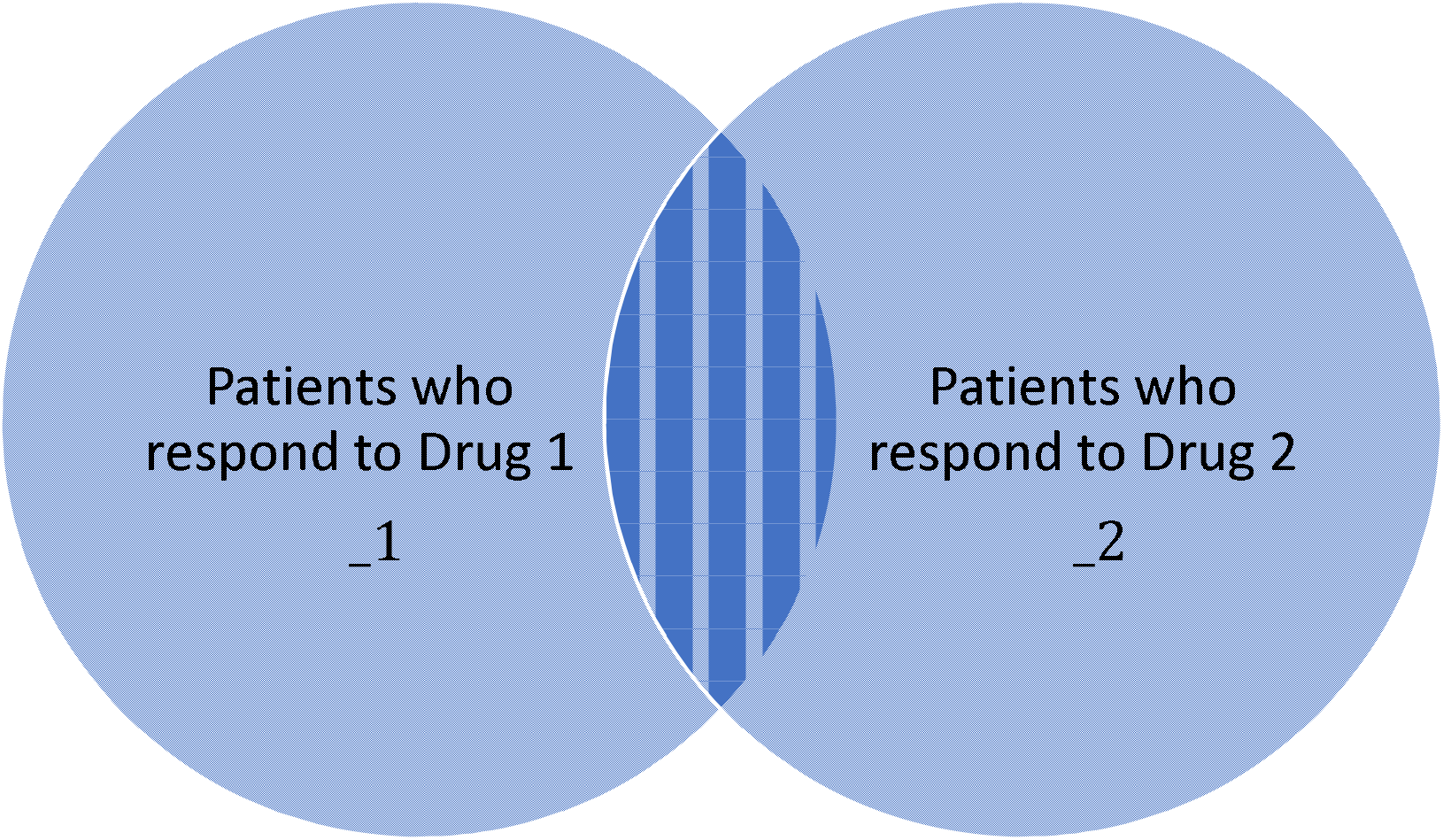
ORR based on the Bliss independence model. The overall area represents the patients who may respond to the combination therapy, and the overlapping area represents those who can potentially respond to both drug components. The reduction of ORR after the combination as compared to the sum of individual ORRs is.

**Figure 3.**
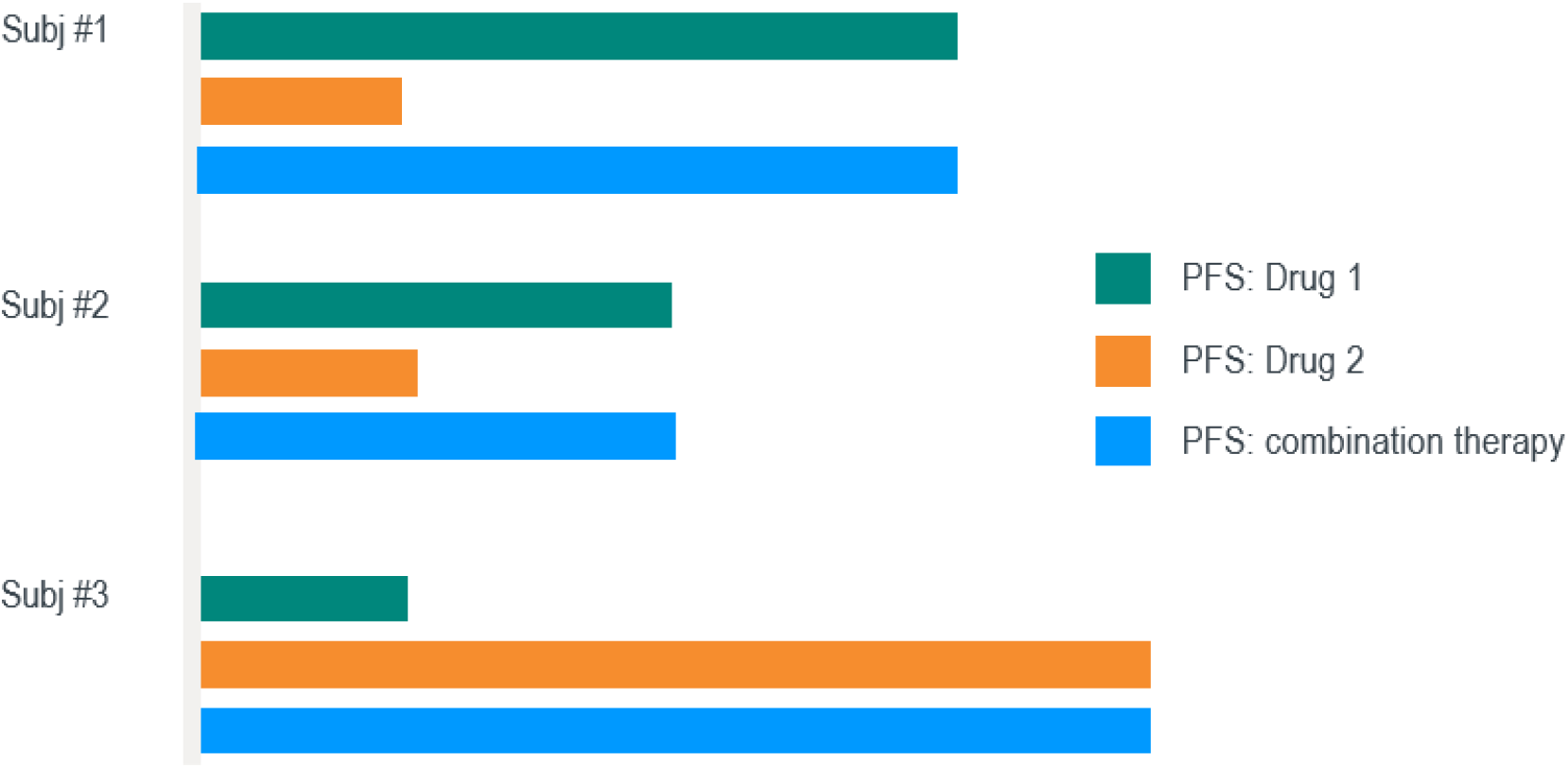
PFS duration based on the independent drug action model. The PFS duration for each patient after treatment with a combination therapy is the longer one of those from the drug components. The first two patients benefit more from Drug 1 and the third patient benefits more from Drug 2.

## Results

Analysis of MRD involves both ORR and DoR. DoR may be different among the following three groups of patients: patient who potentially respond to Drug 1 but not Drug 2, patients who potentially respond to Drug 2 but not Drug 1, and patients who potentially respond to both drugs. For a patient who potentially responds to both drugs, based on the independent drug action model, the DoR is driven by the drug component to which the patient’s tumor is most sensitive and can be predicted from a similar analysis as for PFS. At the aggregate level, the DoR in this group is longer than the other two. Although we cannot differentiate patients among the three groups in practice, the independent action model allows us to account for the difference in the prediction of the DoR for the combination therapy. Notice that, consistent with clinical trial data, the predicted DoR of a combination therapy generally fall between the two individual components^13^.

Let *d*_1_ and *d*_2_ be the respective DoRs of the two drug components, and *p*_1_ and *p*_2_ be the respective ORRs. The MRD of Drug 2 is *d*_2_*p*_2_, but is expected to add less to the combination therapy, just like ORR and PFS. After extending the methods for ORR and PFS to MRD, we arrive at the following index:

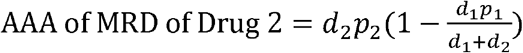

Clearly, any new drug with single-agent ORR too low (or DoR too short) is unlikely to add much antitumor activity to the combination. This point is often under-appreciated in developing innovative immunotherapies, which may partially explain why successes in the field have been rare and infrequent so far despite tremendous effort.

Further inspection of the index reveals that DoR of the new drug has higher impact on the antitumor activity of the combination therapy than ORR. This means that when two drugs have equivalent MRD (e.g., one with 40% ORR and 6-months of DoR and the other with 20% ORR and 12-months of DoR), the one with lower ORR but longer DoR is preferred and may have a better chance to be demonstrated in a clinical trial as long as there is adequate duration of follow-up. A more careful inspection of the index reveals that AAA of Drug 2 is more impacted by ORR than by DoR of Drug 1. This means that there is more room for improvement over SOC with lower ORR (but longer DoR) than over SOC with shorter DoR (but higher ORR).

The DoR can be drastically different among drugs with different mechanisms of action. When Drug 2 has a DoR far exceeding Drug 1, the AAA is close to *d*_2_*p*_2_ (i.e., MRD of Drug 2 is entirely retained after combination). In the metastatic setting, immune checkpoint inhibitors are known to be associated with longer DoR than chemotherapy. This explains why combinations of some immune checkpoint inhibitors with chemotherapy greatly improves antitumor activity compared to chemotherapy alone (e.g., pembrolizumab with pemetrexed-platinum in 1L non-squamous NSCLC in Keynote-189^15^). Conversely, When Drug 1 has a DoR far exceeding Drug 2, the AAA of Drug 2 is close to *d*_2_*p*_2_(1 −*p*_1_). It is solely derived from the proportion of patients who do not respond to Drug 1, predicting the challenges in developing new drugs with short DoR for combining with established immune checkpoint inhibitors, particularly in cases where monotherapy ORR to checkpoint inhibitors is high.

We now apply the index to the development of new drugs for combination with pembrolizumab and pemetrexed-platinum in 1L non-squamous NSCLC. The ORR was 48.0% and the median DoR was 12.4 months for the SOC established after Keynote-189. The MRD is therefore ∼6 months (i.e., 0.48*12.4). Assuming that the OS benefit of a new drug in combination with the SOC can be reasonably predicted from Figure 1 (middle right), it needs to improve the MRD by ∼3 months (i.e., from 6 months to 9 months) to have a clinically meaningful survival improvement of 4-5 months. This means that the new drug combination needs to have a median DoR of ∼15 months when it has a 60% ORR or have a median DoR of ∼12 months when it has a 75% ORR. To get a sense of the required monotherapy activity for the new drug, based on the AAA index of MRD, a new drug with monotherapy ORR of 20% needs to have a median DoR of ∼19 months, which is potentially achievable for a novel immunotherapy with moderate ORR but long DoR. A new drug with a high monotherapy ORR of 40% and moderate DoR of 10 months may also be adequate, which is potentially achievable for an antibody drug-conjugate, KRAS inhibitor or a potent non-immunotherapy. However, drugs with less monotherapy activity may have a hard time to improve the MRD by 3 months.

## Discussion

We have derived three simple indexes for ORR, PFS and MRD for practitioners to use for assessing the AAA of a new drug when combined with any other drug. Examination of the indexes provides important insight into oncology drug discovery and development. The index for MRD is especially relevant to drug development in the immune-oncology field given its high predictive value of long-term clinical benefit. The three indexes are all built upon the independent drug action model under the assumption that there is little cross-resistance or collateral sensitivity between the drug components. Caution must be exerted when violation of the assumption is strongly suspected.

DoR is an integral part of MRD. While routinely reported in practice, median DoR is unreliable when the number of responders is small and is vulnerable to tumor assessment frequency. Therefore, it should be interpreted with caution. Often monotherapy data of a new drug is not available in the first line setting but is available in later lines. As appropriate, monotherapy data in first line for drugs of the same class or historical relationships in monotherapy activity between first line and later lines for drugs in similar classes may be used to assist with the predictions of first line data. Be it based on direct estimation or indirect prediction; the underlying uncertainty should always be properly considered when applying the AAA index of MRD to clinical development decisions.

The long-term clinical benefit of a combination therapy may be confounded with comorbidity, toxicities from the combination that result in high rates of dose reduction and/or study withdrawal, and subsequent therapies post progression, and therefore may not perfectly correlate with any of the index values. Similarly, while clinical benefit from durable stable disease can be significant, it is not incorporated into our analyses. Finally, given the increasing importance of DoR in oncology drug development, a concerted effort for trial Sponsors to routinely report not only the medians but also the relevant Kaplan-Meier plots will benefit all involved parties as Kaplan-Meier plots are more informative than medians even under limited follow-up.

## Data Availability

All data produced in the present work are contained in the manuscript

## Acknowledgement

All authors are stockholders of pharmaceutical companies and can potentially benefit from the publication of this research work.

## Supplementary Appendix

### 1. Phase 3 clinical trials and data used for generating Figure 1

The following data (Table 1) were taken from the primary publications of the referenced studies in US labels. Trials with missing median DoRs (primarily due to inadequate follow-up) are excluded (median DoRs in Keynote-006 and Checkmate-067, the two first Phase 3 studies of immune checkpoint inhibitors in melanoma, are still only partially available in the latest respective 5-year and 6.5-year update). For trials with two experimental arms, to avoid over representation, only the ones approved in the label are excluded. Trials in renal cell carcinoma stand out with long OS.

**Table 1.**
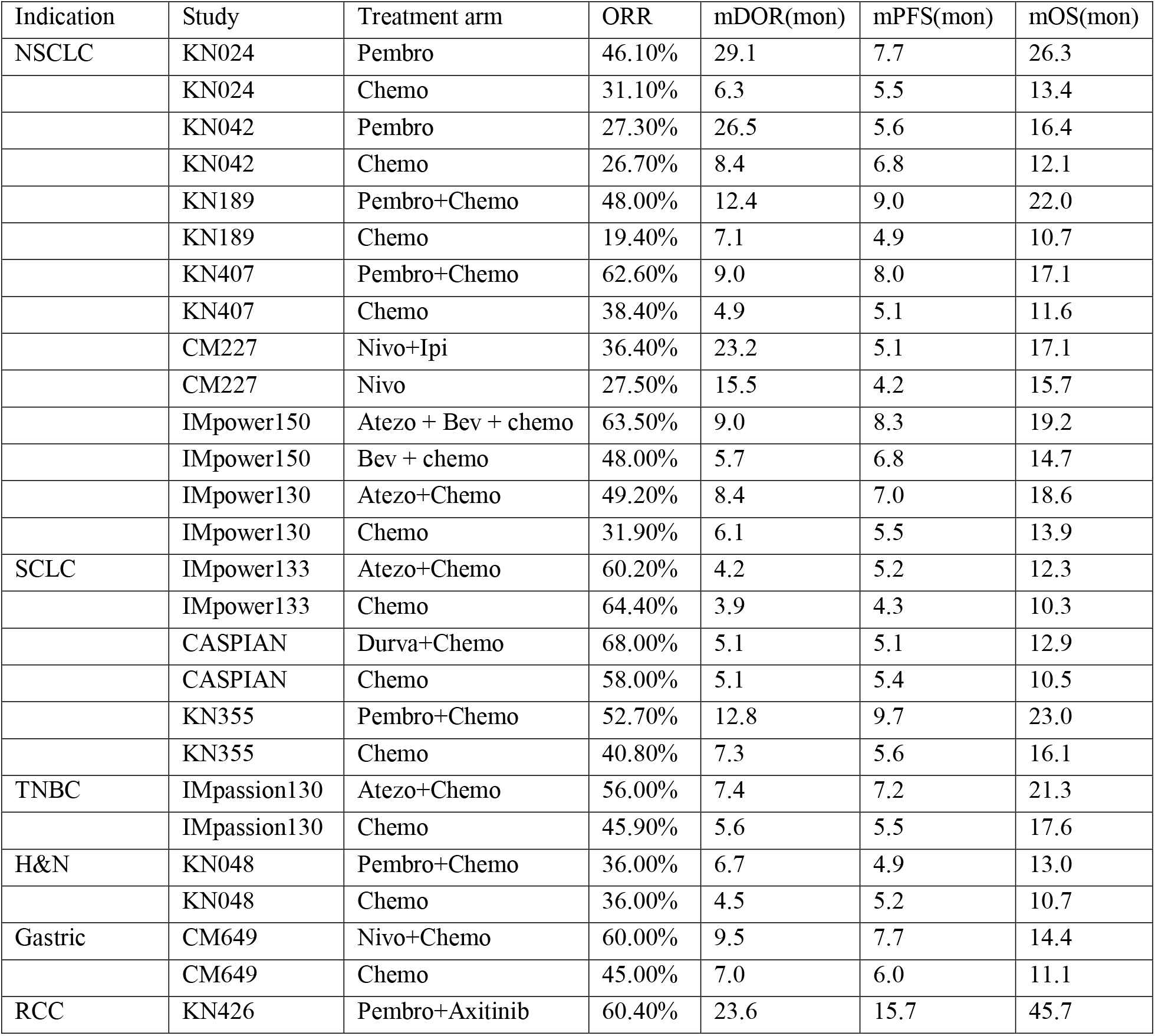
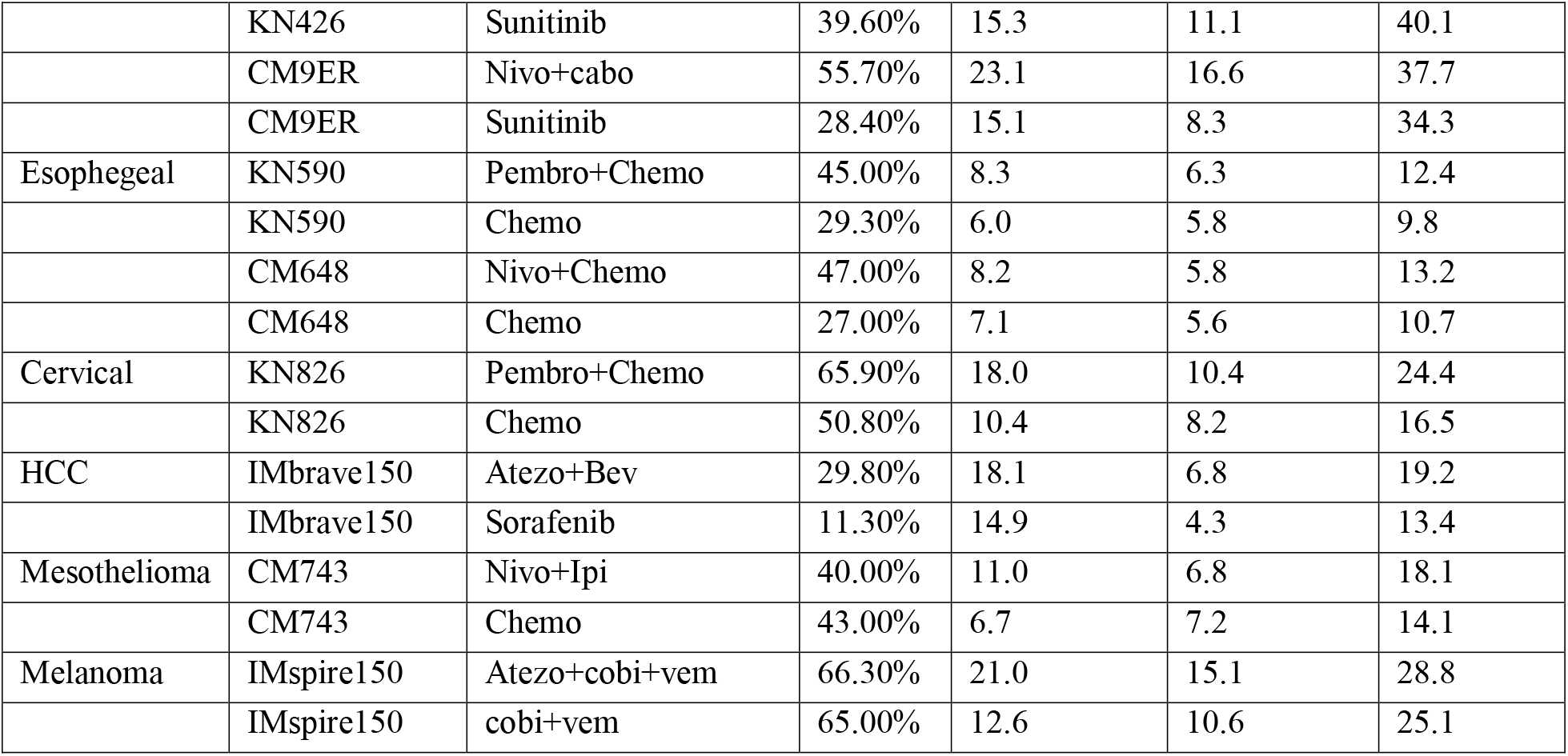
ORR, median DoR (mDOR), median PFS (mPFS) and median OS (mOS) data from the confirmatory Phase 3 trials of anti-PD-(L)1 immune checkpoint inhibitors referenced in US labels

### 2. AAA of PFS

Let *T*_1_ and *T*_2_ be the random variables for PFS of the two drugs, and *T*_12_ be the random variable for PFS of the combination therapy. Based on the independent drug action model, the PFS for a patient is driven by the drug component to which his or her tumor is most sensitive (i.e., *T*_12_=max {*T*_1_, *T*_2_}. Let *t*_1_, *t*_2_, *t*_12_ be the expected (or mean) value of *T*_1_, *T*_2_, *T*_12_, respectively.

When *T*_*i*_ follows an independent exponential distribution (i.e., disease progression occurs continuously and independently at a constant rate over time) with mean 1/ *λ*_*i*_, the survival functions for PFS of the combination therapy *S*(*t*) and the survival functions of PFS for the individual drugs *S*_*i*_(*t*) (*i* = 1,2) have the following relationship:

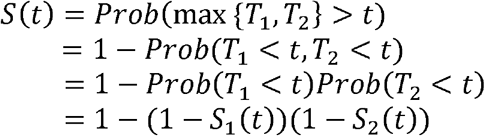

Therefore the density function for *T*_12_ is *f*(*t*) = *λ*_1_*S*_1_ (*t*) + *λ*_2_*S*_2_(*t*) − (*λ*_1_ + *λ*_2_)*S*_1_(*t*)*S*_2_(*t*), and 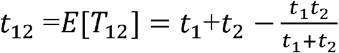. The AAA of PFS from Drug 2 is 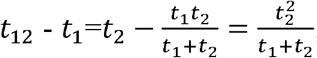 the reduction of PFS after combination as compared to the sum of individual PFS is 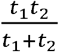 which is equal to half the harmonic mean of *t*_1_ and *t*_2_. In practice, the estimation of mean PFS duration is often restricted with trial follow-up time. Because the ratio between mean and median is fixed under the exponential distribution assumption, mean PFS may be replaced with median PFS for comparing the relative AAA between different drugs.

### 3. AAA of MRD

We start with the ORR analysis. In general, ORR based on the independent drug action model can be best presented in the following classical 2×2 table (Table 2), where 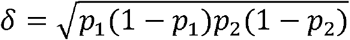 represents the mathematical product of standard deviations of the individual ORRs and *ρ* represents the correlation between individual patient’s responses to Drug 1 and Drug 2. A positive correlation coefficient is an indication of cross-resistance between the two drugs, and a negative correlation coefficient may suggest collateral sensitivity between the two. Based on Table 2, AAA of ORR for Drug 2 is *p*_2_ − *p*_1_ *p*_2_ − *ρ δ*, which decreases linearly as *ρ* increases, as would be expected.

**Table 2.** Probabilities of response to the combination therapy under independent drug action model

|  |  | Potentially respond to Drug 1 |  |
| --- | --- | --- | --- |
|  |  | No | Yes |
| Potentially respond to Drug 2 | Yes | $p_2 - p_1p_2 - \rho\delta$ | $p_1p_2 + \rho\delta$ |
| | No | $1 - p_2 - p_1 + p_1p_2 + \rho\delta$ | $p_1 - p_1p_2 - \rho\delta$ |

Let *D*_1_ and *D*_2_ be the random variables for DoR of the two drugs, and *D*_12_ be the random variable for DoR of the combination therapy in those who potentially respond to both drugs. Based on a similar independent drug action model to PFS^12^, the DoR for a patient who can potentially respond to both drug components is driven by the one to which his or her tumor is most sensitive (i.e., *D*_12_ =max {*D*_1_, *D*_2_ }. Let *d*_1_, *d*_2_, *d*_12_ be the expected value (or mean) of *D*_1_, *D*_2_, *D*_12_, respectively. In general, the following inequalities always hold regardless of the underlying distributions:

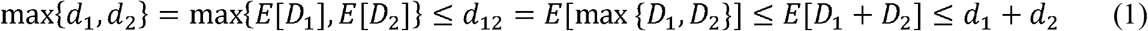

The MRD is *p*_1_ *d*_1_ for Drug 1, *p*_2_*d*_2_ for Drug 2, and (*p*_2_ − *p*_1_ *p*_2_ − *ρ δ*)*d*_1_ (*p*_1_ − *p*_1_ *p*_2_ − *ρ δ*) *d*_2_ + (*p*_1_*p*_2_ +*ρ δ*)*d*_12_ for the combination therapy based on Table 1. Therefore, the AAA of MRD from Drug 2 is the following:

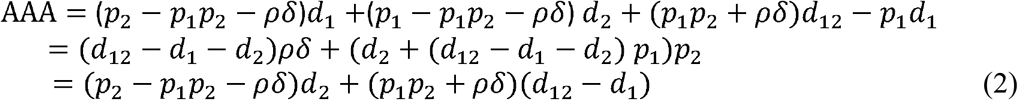

The main result presented in the text is derived under the independent exponential distribution assumption for DoRs and when *ρ* = 0. In this case, the MRD for the combination therapy is 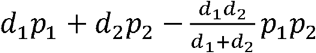 after adopting the formula of *t*_12_ for PFS to *d*_12_, the AAA of MRD from Drug 2 is 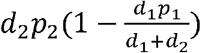, and reduction in MRD after combination as compared to the sum of individual activities is 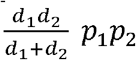. Same as in the PFS analysis, for practical purposes, mean DoR may be substituted with median DoR (or restricted mean) in comparison of the relative AAA of MRD between different drugs.

We now inspect the mathematical property of AAA (eqn. 2) without imposing any distributional assumptions. We start with *ρ* first. Because *d*_12_ − *d*_1_ − *d*_2_ ≤ 0 (eq. 1), the first term in second equality of (eqn. 2) shows that AAA decreases linearly with the cross-resistance level. However, the actual decrease is likely more than linear because *d*_12_ may also decrease as cross-resistance increases. We now focus on measures of individual antitumor activities. The second term in the second equality shows that AAA linearly increases with *p*_2_ since *d*_2_ (*d*_12_ − *d*_1_ − *d*_2_) *p*_1_ ≥ 0 by noticing that it is non-negative even when *p*_1_=1. The first term in the third equality shows that AAA increases linearly with *d*_2_. However, since *d*_12_ is expected to increase with *d*_2_, the second term in the third equality indicates that the increase of AAA with *d*_2_ is likely faster than linear. Because *d*_12_ *≤ d*_1_ + *d*_2_, it is expected from the second term in the second equality that AAA decreases linearly as *p*_1_ increases. From the second term in the third equality, AAA decreases linearly as *d*_1_ increases but the size of decrease is likely offset by an increase of *d*_12_ due to the increase in *d*_1_. Overall, direction-wise, the relationship between AAA of MRD and the measures of individual antitumor activities under the general conditions is consistent with that presented in the main text.

